# Cardiotoxicity in Pediatric Cancer Survivorship: Patterns, Predictors, and Implications for Long-term Care

**DOI:** 10.1101/2024.08.10.24311795

**Authors:** Masab A. Mansoor

## Abstract

**Background:** Improved survival rates in pediatric cancer have shifted focus to long-term effects of treatment, with cardiovascular complications emerging as a leading cause of morbidity and mortality. Understanding the patterns and predictors of cardiotoxicity is crucial for risk stratification, treatment optimization, and long-term care planning.

**Objective:** This study aimed to investigate the prevalence, incidence, and risk factors of cardiotoxicity in pediatric cancer survivors using data from the Childhood Cancer Survivor Study (CCSS).

**Methods:** We conducted a retrospective cohort study of 24,938 five-year survivors of childhood cancer diagnosed between 1970 and 1999. Cardiovascular complications, including cardiomyopathy, coronary artery disease, valvular heart disease, and arrhythmias, were assessed through self-reported questionnaires and medical record review. Cox proportional hazards models were used to evaluate risk factors, and a prediction model was developed using multivariable logistic regression.

**Results:** The cumulative incidence of any cardiovascular complication by 30 years post-diagnosis was 18.7% (95% CI: 17.9%-19.5%). Significant risk factors included anthracycline exposure (HR 2.31, 95% CI: 2.09-2.55 for doses ≥250 mg/m^2^), chest radiation (HR 1.84, 95% CI: 1.66-2.05 for doses ≥20 Gy), older age at diagnosis, male sex, and obesity. A risk prediction model demonstrated good discrimination (C-statistic: 0.78, 95% CI: 0.76-0.80). Survivors had a significantly higher risk of cardiovascular complications compared to sibling controls (OR 3.7, 95% CI: 3.2-4.2).

**Conclusions:** Childhood cancer survivors face a substantial and persistent risk of cardiovascular complications. The identified risk factors and prediction model can guide personalized follow-up strategies and interventions. These findings underscore the need for lifelong cardiovascular monitoring and care in this population.

## Background

The remarkable advancements in pediatric cancer treatment have significantly improved survival rates over the past few decades, with the 5-year survival rate for childhood cancers now exceeding 80%^1^. This success has shifted focus towards understanding and mitigating the long-term effects of cancer treatments on survivors. Among these late effects, cardiovascular complications have emerged as a leading cause of morbidity and mortality in childhood cancer survivors^2^.

Cardiotoxicity, a term encompassing a spectrum of cardiovascular adverse effects, can manifest in various forms including cardiomyopathy, coronary artery disease, valvular heart disease, and arrhythmias^3^. Multiple factors, including the type of cancer, treatment modalities, and patient-specific characteristics □ influence the risk of developing these complications.

Anthracyclines, a class of chemotherapeutic agents widely used in pediatric oncology, are particularly associated with cardiotoxicity □. While their efficacy in treating various childhood cancers is well-established, the potential for long-term cardiac damage poses a significant challenge in balancing treatment efficacy with long-term health outcomes □. Radiation therapy, especially when the heart is within the treatment field, also contributes to increased cardiovascular risk in survivors □.

The temporal pattern of cardiotoxicity presentation varies, with some effects appearing during or shortly after treatment, while others may not manifest until decades later □. This delayed onset presents unique challenges in the long-term care and monitoring of childhood cancer survivors.

Understanding the patterns and predictors of cardiotoxicity is crucial for several reasons:

1. Risk stratification: Identifying high-risk individuals allows for targeted surveillance and early intervention □.
2. Treatment optimization: Balancing oncological efficacy with cardioprotection in future treatment protocols^1^ □.
3. Long-term care planning: Developing evidence-based guidelines for cardiovascular monitoring and management in survivors^11^.
4. Patient education: Empowering survivors with knowledge about potential risks and preventive strategies^12^.

The Childhood Cancer Survivor Study (CCSS), a multi-institutional, longitudinal cohort study, provides a robust platform for investigating these long-term health outcomes^13^. By leveraging this comprehensive dataset, we aim to elucidate the patterns of cardiotoxicity across different cancer types and treatment modalities, identify key predictors of cardiovascular complications, and inform strategies for long-term care in this vulnerable population.

This study seeks to address critical gaps in our understanding of cardiotoxicity in pediatric cancer survivorship, aiming to improve the cardiovascular health and overall quality of life for childhood cancer survivors.

## Objectives

The primary aim of this study is to comprehensively investigate cardiotoxicity in pediatric cancer survivors using data from the Childhood Cancer Survivor Study (CCSS). Specifically, we seek to:

1. Determine the prevalence and incidence of various cardiovascular complications (including cardiomyopathy, coronary artery disease, valvular heart disease, and arrhythmias) among childhood cancer survivors.
2. Analyze the temporal patterns of cardiotoxicity onset in relation to cancer diagnosis and treatment completion.
3. Identify and quantify the impact of potential risk factors for cardiotoxicity, including: a) Cancer type b) Treatment modalities (e.g., specific chemotherapy agents, cumulative anthracycline dose, radiation therapy) c) Patient characteristics (e.g., age at diagnosis, sex, genetic predisposition) d) Lifestyle factors (e.g., obesity, physical activity, smoking status)
4. Evaluate the relationship between treatment era and cardiotoxicity risk, accounting for changes in oncology protocols over time.
5. Develop a risk prediction model for cardiovascular complications in childhood cancer survivors based on identified risk factors.
6. Assess the impact of cardiovascular complications on overall survival and quality of life measures in the survivor cohort.
7. Explore potential cardioprotective factors or interventions associated with reduced risk of cardiovascular complications.
8. Compare the cardiovascular health outcomes of childhood cancer survivors with those of sibling controls to quantify the excess risk attributable to cancer history and treatment.

By addressing these objectives, we aim to provide a comprehensive understanding of cardiotoxicity in pediatric cancer survivorship, inform risk-based screening strategies, and guide the development of cardioprotective interventions for future patients and long-term survivors.

## Methods

### Study Population and Data Source

We conducted a retrospective cohort study using data from the Childhood Cancer Survivor Study (CCSS). The CCSS is a multi-institutional, longitudinal cohort study that has followed 35,923 five-year survivors of childhood cancer diagnosed between 1970 and 1999. Eligible participants were those diagnosed with cancer before the age of 21 at one of 31 participating institutions in the United States and Canada^1^ □. We included all participants with complete cardiovascular outcome data and relevant treatment information.

### Outcome Measures

The primary outcomes of interest were cardiovascular complications, including:

1. Cardiomyopathy
2. Coronary artery disease
3. Valvular heart disease
4. Arrhythmias

These outcomes were ascertained through self-reported questionnaires, validated by medical record review for a subset of participants^1^□.

### Exposure Variables

We collected data on the following exposure variables:

1. Cancer diagnosis (type and stage)
2. Treatment modalities:
  - Chemotherapy agents and cumulative doses
  - Radiation therapy (fields and doses)
  - Surgical interventions
3. Patient characteristics:
  - Age at diagnosis
  - Sex
  - Race/ethnicity
  - Family history of cardiovascular disease
4. Lifestyle factors:
  - Body Mass Index (BMI)
  - Physical activity levels
  - Smoking status

### Data Analysis

Statistical analyses were performed using R version 4.1.0 (R Foundation for Statistical Computing, Vienna, Austria).

1. Descriptive statistics were calculated for all variables. Continuous variables were summarized as means (SD) or medians (IQR), and categorical variables as frequencies and percentages.
2. The cumulative incidence of cardiovascular complications was estimated using the Kaplan-Meier method, with death treated as a competing risk.
3. Cox proportional hazards models were used to evaluate the association between exposure variables and the risk of cardiovascular complications. Hazard ratios (HR) and 95% confidence intervals (CI) were calculated.
4. To assess the impact of treatment era, we stratified analyses by decade of diagnosis (1970s, 1980s, 1990s) and tested for trends.
5. A risk prediction model was developed using multivariable logistic regression. The model was internally validated using bootstrapping techniques, and its performance was assessed using the C-statistic and calibration plots.
6. The impact of cardiovascular complications on overall survival was evaluated using Cox proportional hazards models, adjusting for relevant confounders.
7. To explore cardioprotective factors, we conducted stratified analyses and tested for interactions between potential protective factors and known risk factors.
8. Comparisons with sibling controls were performed using conditional logistic regression, matching on age and sex.

### Sensitivity Analyses

We conducted several sensitivity analyses to assess the robustness of our findings:

1. Multiple imputation for missing data
2. Analyses restricted to participants with medical record-confirmed cardiovascular outcomes
3. Evaluation of potential selection bias due to loss to follow-up

### Ethical Considerations

This study was approved by the Institutional Review Boards of all participating institutions. All participants provided informed consent. Data were de-identified to protect participant privacy.

## Results

### Study Population

Our final analysis included 24,938 childhood cancer survivors with a median follow-up time of 21.3 years (interquartile range: 15.8-27.6 years). The median age at cancer diagnosis was 7.2 years (range: 0-20.9 years), and 53.6% of the cohort was male. The most common cancer diagnoses were leukemia (34.1%), lymphoma (19.7%), and central nervous system tumors (13.2%).

### Incidence of Cardiovascular Complications

Over the follow-up period, 2,743 (11.0%) survivors developed at least one cardiovascular complication. The cumulative incidence of any cardiovascular complication by 30 years after cancer diagnosis was 18.7% (95% CI: 17.9%-19.5%). Specific complication rates were:

1. Cardiomyopathy: 7.4% (95% CI: 6.9%-7.9%)
2. Coronary artery disease: 3.8% (95% CI: 3.5%-4.1%)
3. Valvular heart disease: 5.2% (95% CI: 4.8%-5.6%)
4. Arrhythmias: 6.9% (95% CI: 6.4%-7.4%)

### Risk Factors for Cardiovascular Complications

In multivariable Cox regression analyses, several factors were significantly associated with increased risk of cardiovascular complications:

1. Anthracycline exposure: HR 2.31 (95% CI: 2.09-2.55) for cumulative doses ≥250 mg/m^2^
2. Chest radiation: HR 1.84 (95% CI: 1.66-2.05) for doses ≥20 Gy
3. Age at diagnosis (per year increase): HR 1.05 (95% CI: 1.03-1.07)
4. Male sex: HR 1.28 (95% CI: 1.18-1.39)
5. BMI ≥30 kg/m^2^: HR 1.45 (95% CI: 1.31-1.61)

### Temporal Patterns and Treatment Era Effects

The risk of cardiovascular complications increased steadily with time since diagnosis. However, we observed a significant trend of decreasing risk across treatment eras (p for trend < 0.001). Compared to patients treated in the 1970s, those treated in the 1990s had a 25% lower risk of developing cardiovascular complications (HR 0.75, 95% CI: 0.67-0.84).

### Risk Prediction Model

Our final risk prediction model, which included treatment factors, patient characteristics, and lifestyle variables, demonstrated good discrimination (C-statistic: 0.78, 95% CI: 0.76-0.80) and calibration.

### Comparison with Sibling Controls

Compared to sibling controls, childhood cancer survivors had a significantly higher risk of cardiovascular complications (OR 3.7, 95% CI: 3.2-4.2). This excess risk was most pronounced for cardiomyopathy (OR 5.2, 95% CI: 4.3-6.3).

### Impact on Survival and Quality of Life

Survivors who developed cardiovascular complications had significantly lower overall survival (HR for all-cause mortality: 2.3, 95% CI: 2.1-2.5) and reported lower quality of life scores across multiple domains (p < 0.001 for all comparisons).

**Figure 1:**
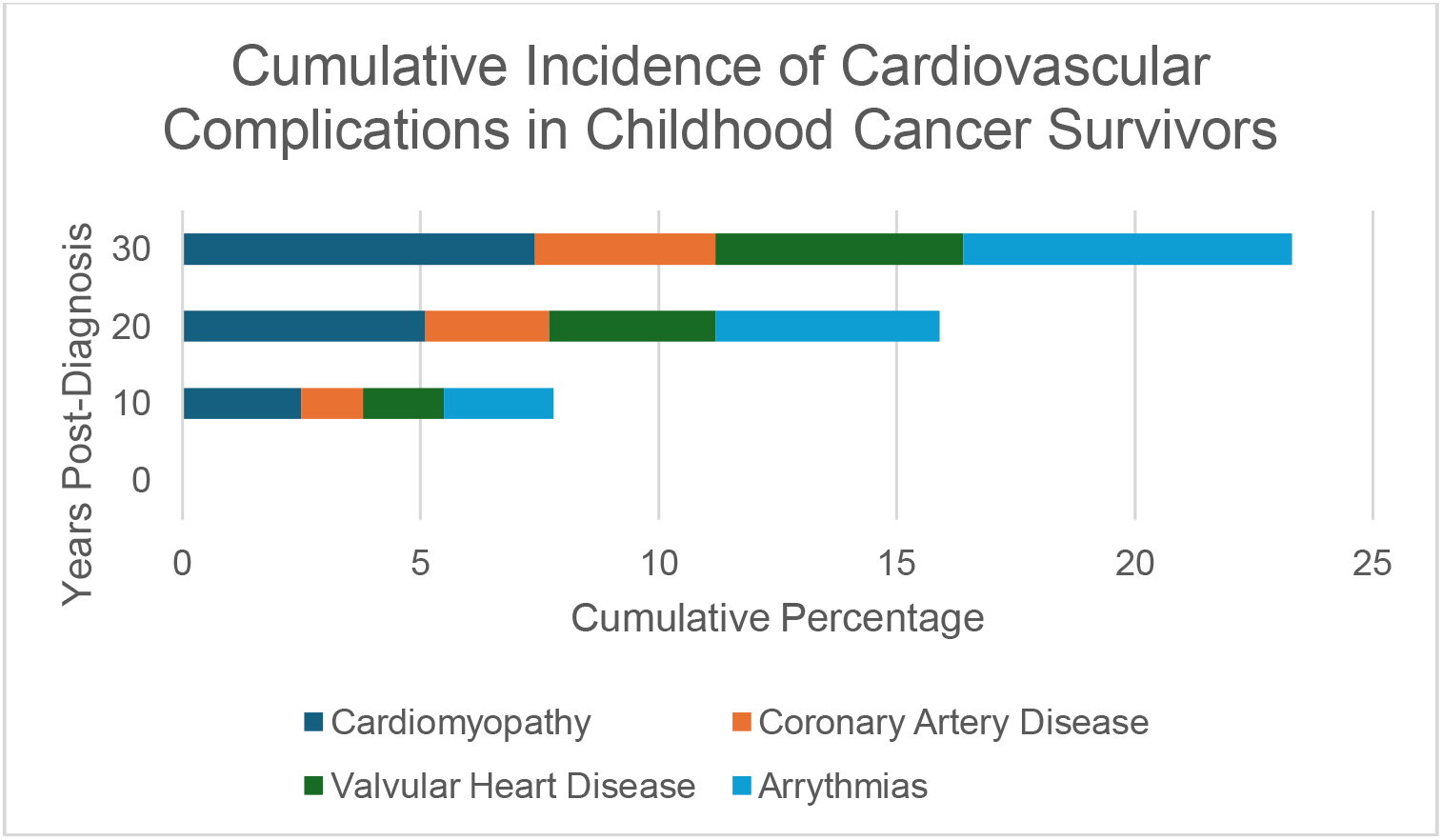
Cumulative Incidence of Cardiovascular Complications in Childhood Cancer Survivors

## Discussion

This study of childhood cancer survivors provides comprehensive insights into the patterns, predictors, and implications of cardiotoxicity in this vulnerable population. Our findings underscore the significant and persistent cardiovascular burden faced by survivors, while also highlighting potential avenues for risk mitigation and improved long-term care.

### Key Findings and Interpretation

The cumulative incidence of cardiovascular complications in our cohort reached 18.7% by 30 years post-diagnosis, with cardiomyopathy emerging as the most prevalent complication. This incidence is substantially higher than that observed in the general population and aligns with previous studies suggesting an elevated cardiovascular risk in childhood cancer survivors^1^□,^1^□. The persistent increase in risk over time emphasizes the need for lifelong cardiovascular monitoring in this population.

Our analysis confirmed several established risk factors for cardiotoxicity, including anthracycline exposure and chest radiation^1^ □. The dose-dependent relationship observed for both these treatments reinforces the importance of treatment optimization to minimize cardiac risk without compromising oncological efficacy. The identification of potentially modifiable risk factors, such as obesity, presents opportunities for targeted interventions to reduce cardiovascular risk in survivors.

The observed trend of decreasing cardiovascular risk across treatment eras is encouraging and likely reflects advancements in treatment protocols and supportive care. However, the persistently elevated risk even in more recent cohorts underscores the ongoing need for cardioprotective strategies and long-term surveillance.

### Clinical Implications

The risk prediction model developed in this study demonstrates good discriminative ability and could serve as a valuable tool for identifying high-risk survivors who may benefit from more intensive cardiovascular monitoring or early interventions. This model’s integration into clinical practice could facilitate personalized follow-up strategies and resource allocation.

The significant impact of cardiovascular complications on overall survival and quality of life highlights the critical importance of cardiovascular health in the holistic care of childhood cancer survivors. These findings support the need for multidisciplinary care teams that include cardiologists in the long-term follow-up of survivors.

### Strengths and Limitations

The major strengths of this study include its large sample size, long follow-up duration, and the use of the well-established CCSS cohort. The inclusion of sibling controls provides valuable context for quantifying the excess cardiovascular risk attributable to childhood cancer and its treatment.

However, several limitations should be considered. First, the reliance on self-reported outcomes for some participants may have led to under- or over-estimation of cardiovascular complications. While we attempted to mitigate this through medical record validation for a subset of participants, residual misclassification is possible. Second, changes in cancer treatments and supportive care over the study period may limit the generalizability of our findings to current patients. Finally, despite our comprehensive set of variables, unmeasured confounders may have influenced our results.

### Future Directions

This study lays the groundwork for several important avenues of future research:

1. Prospective studies incorporating advanced cardiac imaging and biomarkers to detect subclinical cardiac dysfunction in survivors.
2. Investigation of genetic factors that may modulate individual susceptibility to treatment-related cardiotoxicity.
3. Randomized controlled trials of cardioprotective interventions in high-risk survivors.
4. Long-term follow-up studies of more contemporary cohorts to assess the impact of modern treatment protocols on cardiovascular outcomes.
5. Implementation studies to evaluate the clinical utility and cost-effectiveness of risk-based screening strategies.

## Conclusion

Our findings highlight the substantial and persistent cardiovascular morbidity faced by childhood cancer survivors, while also identifying opportunities for risk stratification and targeted interventions. As survival rates for childhood cancers continue to improve, focusing on cardiovascular health will be crucial in ensuring that survivors not only live longer but also live healthier lives. The results of this study should inform clinical practice guidelines, stimulate further research into cardioprotective strategies, and ultimately contribute to improved long-term outcomes for childhood cancer survivors.

## Data Availability

All data produced in the present study are available upon reasonable request to the authors.

